# A Systematic Review of Long-term Antidepressant Outcomes in Comorbid Depression and Type 2 Diabetes

**DOI:** 10.1101/2022.04.11.22273519

**Authors:** Annie Jeffery, Joshua E. J. Buckman, Emma Francis, Kate Walters, Ian C. K. Wong, David Osborn, Joseph F. Hayes

## Abstract

**Background:** Depression is a common and chronic comorbidity affecting approximately one in four people with type 2 diabetes (T2DM), and often lasting several years. Past systematic reviews have been unable to identify evidence for long-term (12+ months) antidepressant treatment outcomes in comorbid depression and type 2 diabetes. These reviews are >10years old, included only randomised controlled trials or had limited search strategies. We aimed to systematically review observational studies for long-term outcomes of antidepressant treatment in adults with comorbid depression and T2DM, including broader, up-to-date searches.

**Methods and findings:** This review was pre-registered on PROSPERO (CRD42020182788). We searched seven databases using terms related to depression, T2DM and antidepressant medication. From 14,389 reports retrieved, 63 were screened at full text stage and 0 met inclusion criteria. The reasons for exclusion at full text stage were: Studies did not meet inclusion criteria for antidepressant treatment (n = 50); studies did not meet inclusion criteria for T2DM (n = 36); studies did not meet inclusion criteria for depression (n = 29); studies did not include follow-up time (n = 25); studies did not meet inclusion criteria for observational study (n = 14); studies did not include any measurable outcomes (n = 5); studies did not include a suitable comparison (n = 3).

**Conclusions:** We found no evidence concerning long-term outcomes of antidepressant treatment in individuals with comorbid depression and T2DM. Insufficient ascertainment of antidepressant prescription, case identification, and short follow-up times are the primary reasons for this. Research is urgently required to determine long-term outcomes associated with antidepressant treatment in this patient group.

## 1. BACKGROUND

It is estimated that one in four people with type 2 diabetes (T2DM) have comorbid depression [1], and there is considerable evidence showing increased prevalence rates of depression in individuals with T2DM compared to those without [2]. Depression has been shown to be associated with poor glycaemic control [3], the development of diabetic complications [4] and decreased adherence to diabetic treatments [5]. Therefore, the successful treatment of depression in individuals with T2DM can be important to managing both physical and mental health.

Antidepressant medication is recommended by national and international healthcare guidelines as a treatment option for people with moderate to severe depression [6]–[9]. However, guidelines addressing the treatment of depression in individuals with physical long-term conditions are limited and non-specific [10]. A review of pharmacological treatment guidelines with respect to multimorbidity notes potential dangers of applying current guidelines to patients with multiple conditions [11]. A number of commonly prescribed antidepressants cause side-effects that potentially exacerbate T2DM or its complications, such as weight gain [12], hypoglycaemia, cardiac complications, arthralgia, gastrointestinal disturbances, sexual dysfunction, and visual impairment [13]. Indeed, selective serotonin re-uptake inhibitors, which are the most commonly prescribed class of antidepressants [6] are cautioned for use in people with diabetes mellitus [13].

A 2012 Cochrane review [14] and an earlier 2010 review [15] examined randomised-controlled trials (RCTs) of psychological and pharmacological interventions for people with depression and T2DM. They found that antidepressant medications improve both depressive symptoms and glycaemic control in the short term. However, the duration of the trials ranged from 6 weeks to 6 months from the start of treatment, and longer-term outcomes are therefore unknown.

Understanding longer-term outcomes of antidepressant use is important in individuals with comorbid depression and T2DM. While initial response to antidepressant treatment is expected in 4-6 weeks [6], depression is often chronic, many patients experience disabling sub-syndromal symptoms for several years, or multiple relapses or recurrences of depression after having initially recovered from an episode [16]–[19]. As a result, it is increasingly common for patients to have antidepressants prescribed for two or more years in order to reduce the risk of relapse, and in many cases this can lead to indefinite treatment with antidepressants [20]–[22]. In the general population, there is evidence of increased risk of relapse or harmful outcomes for those who remain on antidepressants for longer durations [23], [24]. Therefore, understanding the impact of antidepressant treatment over longer follow-up periods is important [25]. Similarly, longer term studies are needed to understand the impact of antidepressant treatment on important long-term clinical outcomes, such as disease stage progression, diabetic complications and mortality.

While RCTs are the gold standard for investigating treatment outcomes, longer follow-up times may not be feasible due to high costs and participant attrition [26]. Observational studies, therefore, may be better suited to finding evidence for long-term outcomes [27].

A 2017 systematic review by Roopan et al included “different study designs”, however, did not specify which study designs were included and they only searched a limited number of databases [28]. This review found one observational study [29], using routinely collected data over a period of 11 years, which found long-term antidepressant treatment (3+ years) to be associated with higher risk of hypoglycaemia. However, it was not a requirement for participants to be diagnosed with depression or present with depressive symptoms, and the reason for prescribing the antidepressants were unknown.

We aimed to systematically review the long-term outcomes of antidepressant treatment in adults with comorbid depression and T2DM. We focused our review on observational studies, as previous investigation of these has been limited.

## 2. METHODS

We prepared and presented this systematic review in accordance with the Preferred Reporting Items for Systematic Reviews and Meta-analysis (PRISMA), and registered the review on PROSPERO prior to the commencement of screening (CRD42020182788).

### 2.1 Study inclusion criteria

#### 2.1.1 Types of study design

We included observational studies from routinely collected data, registry studies, cohort studies, case-control studies and cross-sectional studies. We excluded RCTs, quasi-experimental trials where selected patients receive an antidepressant as part of the study, systematic reviews, case studies/reports, editorials, letters and opinion pieces.

#### 2.1.2 Participants

We included studies investigating adults (18+ years) with comorbid depression and T2DM:

Depression could be identified by clinician diagnosis, medical records, standardised interviews, or self-report. Where diagnostic criteria were not available, we used the authors’ definition of depression, provided depression was explicitly stated for all participants or the subgroup being used for analysis. We did not include studies where depression was defined by the prescription of an antidepressant alone, as there are other indications for antidepressants.

Type 2 diabetes could be defined as an in-study clinician diagnosis, medical record, or self-report. The type of diabetes should have been explicitly verified as type 2, and we did not include studies where the diabetes type was ambiguous or mixed.

We excluded participants with mental disorders other than depression, e.g. bipolar disorder, where the primary diagnosis was not depression.

#### 2.1.3 Intervention

The intervention of interest was any antidepressant medication taken for a minimum duration of 6 months, according to treatment guideline recommendations [6]. We included antidepressant prescriptions defined through self-report, prescription records or clinician report. We excluded studies where the antidepressant was prescribed for a different mental or physical disorder, such as anxiety or neuropathic pain.

#### 2.1.4 Comparison

The comparison group was no antidepressant treatment, either in the same individual or in a control group.

#### 2.1.5 Outcomes

As this review was exploratory, we did not limit the potential outcomes that could be measured in association with antidepressant treatment. These could include: depression severity, remission, relapse or recurrence; glycaemic control or other markers of diabetic health; diabetic comorbidities; other measures of physical or mental health; socio-economic outcomes; quality of life; healthcare utilisation or costs; mortality.

#### 2.1.6 Time

We included studies with a minimum follow-up time of 12 months after commencement of antidepressant treatment.

### 2.2 Search strategy and screening

We used Medical Subject Headings (MeSH) and keywords for: depression AND type 2 diabetes AND antidepressant; to search the following sources for the identification of studies, from inception to 10-May-2021:

- MEDLINE
- EMBASE
- Scopus
- CINAHL Plus
- Web of Science
- PsycInfo
- PsycExtra
- Open Grey

We included articles in the following languages: English, French, Spanish, Italian, Portuguese, Greek.

The first (AJ) and third (EF) authors independently screened all titles and abstracts against the eligibility criteria. The first (AJ) and second (JB) review authors independently carried out full-text screening of studies that potentially met our inclusion criteria.

In addition, we reviewed references of all studies screened at full text stage and all relevant systematic reviews found during the search.

Reasons for exclusion were independently recorded by the first and second authors. Any disagreement was resolved through discussion.

### 2.3 Data collection and analysis

No data collection or analysis was performed in this review due to a lack of available studies after the search, however, the original analysis plans are detailed in the PROSPERO protocol CRD42020182788.

## 3 RESULTS

Our search yielded 14,389 unique abstracts, of which 63 full-texts were assessed for eligibility [30]– [92]. We found no studies that met our inclusion criteria. A flow chart of study exclusion is shown in Figure 1.

**Figure 1.**
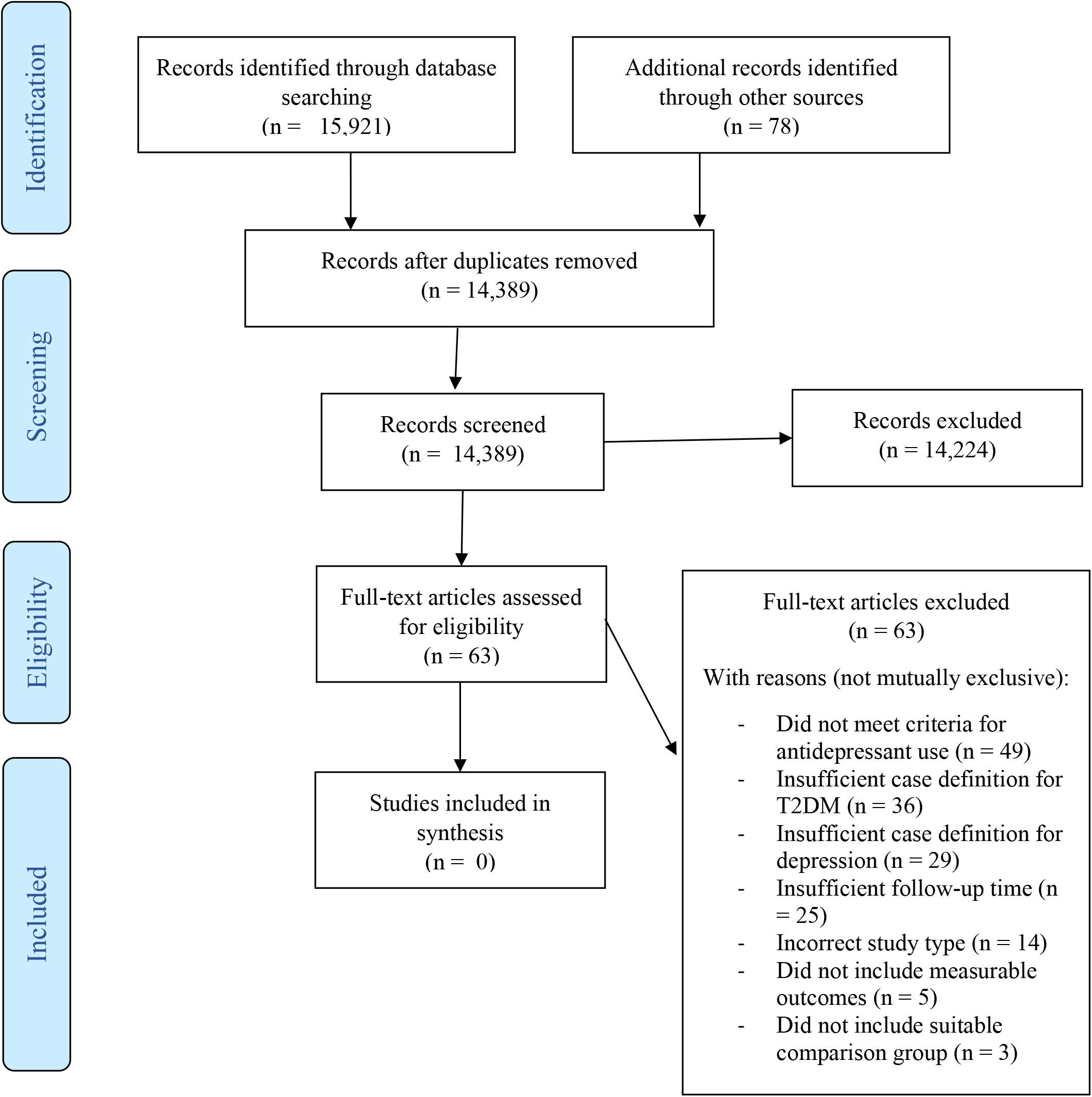
Flowchart of study exclusion *From:* Moher D, Liberati A, Tetzlaff J, Altman DG, The PRISMA Group (2009). *P*referred *R*eporting *I*tems for *S*ystematic Reviews and *M*eta-*A*nalyses: The PRISMA Statement. PLoS Med 6(7): e1000097. doi:10.1371/journal.pmed1000097

### 3.1 Reasons for exclusion

Full reasons for exclusion of all studies excluded at full text screening stage are given in Table 1. The reasons for exclusion were not mutually exclusive – a study could have more than one reason for exclusion.

**Table 1.** Reasons for exclusion at full text screening stage

| Reference | Population |  | Intervention |  | Comparison | Outcome |  | Study Design |
| --- | --- | --- | --- | --- | --- | --- | --- | --- |
|  | Diabetes type 2 specified and distinct | Participant meets criteria for depression | Antidepressant use specified and distinct | Antidepressant treatment duration at least 6 months | Suitable comparison group | Measurable outcome | Outcome measured at least 12 months after start of treatment | Observational study |
| Abraham 2009 (30) | Y | Y | Y | Y | Y | Y | N | N |
| Abraham 2012 (31) | Y | Y | Y | Y | Y | Y | N | N |
| Acee 2012 (32) | Y | N | Y | N | Y | Y | N | Y |
| Alenzi 2016 (33) | N | Y | Y | N | Y | Y | Y | Y |
| Amital 2012 (34) | N | Y | Y | N | N | N | NA | Y |
| Arya 2017 (35) | N | Y | Y | N | Y | Y | Y | Y |
| Battacharya 2016 (36) | Y | Y | Y | N | Y | Y | Y | Y |
| Bryan 2009 (37) | N | Y | Y | N | Y | Y | N | N |
| Caughey 2013 (38) | N | N | Y | N | Y | Y | N | Y |
| Danhauer 2018 (40) | Y | N | Y | N | Y | Y | N | Y |
| Derijks 2008A (41) | N | N | Y | Y | Y | Y | Y | Y |
| Derijks 2008B (42) | N | N | Y | Y | Y | Y | Y | Y |
| Filipic 2010 (43) | Y | Y | Y | Y | Y | Y | N | N |
| Hazuda 2019 (44) | Y | N | Y | N | Y | Y | Y | Y |
| Higgins 2006 (45) | Y | Y | Y | N | Y | Y | N | Y |
| Ioscifescu 2004 (46) | N | Y | Y | Y | N | Y | N | N |
| Kammer 2015 (47) | N | N | Y | N | Y | Y | N | Y |
| Kammer 2016A (48) | N | N | Y | N | Y | Y | N | Y |
| Kammer 2016B (49) | N | Y | Y | N | Y | Y | N | Y |
| Katon 2008A (50) | NA | NA | NA | NA | NA | NA | NA | N |
| Katon 2008B (51) | N | Y | N | NA | NA | Y | NA | N |
| Katon 2011 (52) | NA | NA | NA | NA | NA | NA | NA | N |
| Katon 2012 (53) | Y | N | N | NA | NA | Y | NA | Y |
| Katon 2015 (54) | N | N | N | NA | NA | Y | NA | Y |
| Keating 2018 (55) | Y | N | N | NA | NA | Y | NA | Y |
| Keitner 1991 (56) | N | Y | N | NA | NA | Y | NA | Y |
| Knol 2008 (57) | N | N | Y | Y | Y | Y | Y | Y |
| Labad 2012 (58) | Y | N | Y | N | Y | Y | N | Y |
| Lee 2017 (59) | N | N | Y | N | Y | Y | Y | Y |
| Lee 2020 (60) | N | N | Y | N | Y | Y | Y | Y |
| Lin 2009 (61) | N | Y | N | NA | NA | Y | NA | Y |
| Lunghi 2017A (62) | Y | Y | N | NA | NA | Y | NA | Y |
| Lunghi 2017B (63) | Y | Y | N | NA | NA | Y | NA | Y |
| Lustman 1997 (64) | N | Y | Y | N | Y | Y | Y | N |
| Lustman 2007 (65) | Y | Y | Y | Y | Y | Y | N | N |
| Miller 1996 (66) | N | Y | Y | Y | Y | Y | N | N |
| Moulton 2019 (67) | Y | N | N | NA | NA | Y | NA | Y |
| Nicolau 2013 (68) | Y | Y | Y | Y | Y | Y | N | N |
| Noordamn 2016 (69) | Y | N | Y | N | Y | Y | Y | Y |
| Novak 2016 (70) | N | N | N | NA | NA | Y | NA | Y |
| Osborn 2010 (71) | N | Y | Y | N | Y | N | NA | Y |
| Pan 2011 (72) | Y | N | N | NA | NA | Y | NA | Y |
| Passamonti 2015 (72) | Y | Y | Y | N | Y | Y | N | Y |
| Peyrot 1999 (73) | N | Y | N | NA | NA | Y | NA | Y |
| Popkin 1985 (74) | N | Y | Y | N | Y | Y | N | Y |
| Radojkovic 2016 (75) | Y | Y | Y | Y | Y | Y | N | N |
| Rivich 2019 (76) | N | N | Y | N | Y | Y | N | Y |
| Rizvi 2014 (77) | N | Y | Y | N | N | N | NA | Y |
| Rubin 2013 (78) | Y | N | Y | N | Y | Y | Y | Y |
| Rush 2008 (79) | N | Y | Y | N | Y | Y | Y | Y |
| Sambamoorthi 2006 (80) | N | Y | Y | N | Y | N | NA | Y |
| Shen 2013 (81) | N | Y | Y | N | Y | Y | Y | Y |
| Simon 2005 (82) | N | N | Y | N | Y | N | NA | Y |
| Song 2014 (83) | N | Y | Y | Y | Y | Y | N | Y |
| Tatonetti 2011 (84) | N | N | Y | N | Y | Y | N | Y |
| Tuncel 2015 (85) | Y | N | Y | N | Y | Y | N | Y |
| Unutzer 2009 (86) | N | N | N | NA | NA | Y | NA | Y |
| Viscogliosi 2020 (87) | N | N | Y | N | Y | Y | N | Y |
| Wami 2013 (88) | Y | N | Y | N | Y | Y | Y | Y |
| Wickstrom 1964 (89) | N | Y | Y | N | Y | Y | N | N |
| Willems 2010 (90) | N | N | N | NA | NA | Y | NA | Y |
| Xing 2017 (91) | Y | Y | N | NA | Y | Y | NA | Y |
| Yekta 2015 (92) | Y | N | Y | Y | Y | Y | Y | Y |

#### 3.1.1 Study type

Fourteen studies did not meet our definition of an observational study [30], [31], [37], [42], [45], [49]–[51], [63], [65], [67], [75], [89], [93]. Two of these [49], [51] were descriptive narratives and so all other inclusion criteria was marked as “NA”. The remaining studies all included individuals who received antidepressant treatment as part of the study, however, none of these met all remaining requirements for inclusion criteria.

#### 3.1.2 Case definition

There were 36 studies that did not meet inclusion criteria for T2DM. Of these, five studies included T2DM as one of many undistinguished comorbidities [41], [45], [55], [65], [81] and four studies did not distinguish diabetic from non-diabetic participants [35], [46]–[48]. The remaining studies (n = 27) did not adequately differentiate type 2 diabetes from other diabetes types [33], [34], [37], [38], [41], [50], [52], [56], [58]–[60], [63], [69], [70], [73], [74], [76], [77], [79], [80], [82]–[84], [86], [87], [89], [90].

There were 29 that studies did not meet inclusion criteria for the case definition of depression. Of these, six studies included antidepressant treatment in their case definition for depression [52], [53], [69], [71], [86], [90]. Eleven studies specifically included participants who did not have depression [32], [39], [47], [48], [57], [66], [76], [78], [82], [87], [88]. The remaining studies (n = 12) did not specify whether participants had depression [38], [40], [41], [43], [54], [56], [58], [59], [68], [84], [85], [92].

#### 3.1.3 Intervention inclusion criteria

The most common reason for exclusion (n = 49) was the failure to meet our inclusion criteria for antidepressant treatment. There were 15 studies which did not identify a group of participants who were all known to have received antidepressant prescriptions through self-report, prescription records or clinician report [50], [52]–[55], [60]–[62], [66], [69], [71], [73], [86], [90], [91]. Six of these only included antidepressant treatment as a potential case definition for depression, and did not distinguish between participants who did or did not receive antidepressant treatment [52], [53], [69], [71], [86], [90]. Two studies [54], [91] did not explore antidepressant treatment separately from other psychotropics, and one study [50] grouped antidepressant treatment with non-pharmacological treatment for depression.

In addition, there were a further 34 studies in which the duration of antidepressant treatment was not specified or which did not meet our inclusion criteria of a minimum of 6 months duration. Four of these studies measured “any” historic antidepressant use with unspecified durations [38], [77], [78], [84]; three studies specified antidepressant treatment durations of less than 6 months [58], [72], [74]; and two studies measured patterns of antidepressant use as their outcome, again with unspecified durations [34], [82]. The remaining studies were either cross-sectional or measured only current antidepressant use at a single baseline date, without information on the length of antidepressant treatment [32], [33], [35]–[37], [39], [40], [43], [44], [46]–[48], [57], [59], [68], [70], [79]–[81], [83], [85], [87]–[89]

With exception of one study [36], all of the studies that were excluded based on the 6 month duration of antidepressant treatment criteria, had additional reasons for exclusion. Therefore, only one study was excluded on this criteria alone. This study [36] examined the health expenditure in people with comorbid depression and type 2 diabetes, comparing those who received no treatment for depression and those who received antidepressant treatment. The sample contained 5,295 individuals who were Medicare enrolees in the USA, and the results were propensity score adjusted. This study showed that participants treated with antidepressants at baseline had a 16% reduction in all healthcare expenditures across a 12 month period (−0.16, 95% CIs -0.23 to -0.10).

#### 3.1.4 Comparison

There were three studies that did not include a suitable comparison group to individuals using antidepressant [34], [45], [77]. If the study did not distinguish participants who received antidepressant treatment, this inclusion criteria was marked as “NA”.

#### 3.1.5 Outcomes

There were five studies that did not include measurable outcomes of antidepressant treatment [34], [70], [77], [80], [82]. These studies described the prevalence of antidepressant prescribing, and sometimes factors associated with antidepressant prescribing, however, they did not describe subsequent outcomes. If the study did not distinguish participants who received antidepressant treatment, this inclusion criteria was marked as “NA”.

#### 3.1.6 Follow up time

There were 25 studies that did not include our minimum follow-up time of 12 months. Nine studies presented results from cross-sectional analyses [39], [44], [46]–[48], [57], [76], [85], [87]. Two studies were unclear with regards to the timing of the intervention and the outcome [38], [84]. The remaining studies were longitudinal with a maximum follow-up time of 6 months [30]–[32], [37], [42], [45], [46], [56], [64], [65], [67], [71], [74], [75], [83], [89]. If the study did not distinguish participants who received antidepressant treatment, this inclusion criteria was marked as “NA”.

## 4 DISCUSSION

### 4.1 Summary of findings

This systematic review set out to investigate long-term outcomes from observational studies in individuals with comorbid depression and type 2 diabetes prescribed antidepressants. However, no studies matched our inclusion criteria. The main reasons for exclusion were unspecified case definitions for depression and type 2 diabetes, short or unspecified duration of antidepressant treatment and short overall follow-up time to assess outcomes.

The majority of studies focused on participants with diabetes, however, 27 of these did not adequately specify type 2 diabetes. Historically, the classification of diabetes types has been blurred. For example, prior to the implementation of ICD-10, the International Classification of Diseases grouped diabetes codes by complication, with the diabetes type included only as an optional add on [94]. In addition, a report on the coding of diabetes in UK healthcare noted that patients were commonly unaware of their diabetes type [95] – if this is the case, it may also make the identification of diabetes type through self-report challenging or unreliable. Another option to identify diabetes type may be based on assumptions made from the medications prescribed [38]. The introduction of “Type 2” diabetes in 1998, replaced “non-insulin dependent diabetes” [96] – a term which is still used today [97], [98]. However, this terminology may lead to unreliable definitions of diabetes type: Many individuals with T2DM progress to insulin therapy [99], and other diabetes types or conditions may use oral antidiabetic medication such as metformin [100]. Thus, the identification of diabetes type may be challenging in observational studies due to the use of different nomenclature over time.

Similarly, the definition of depression seems to be heterogeneous in observational studies. Studies using data from clinical sources have shown that depression is underdiagnosed generally and especially so among those with comorbid physical health problems [101]–[103]. Indeed, more than one third of the studies screened at full text stage did not specify whether participants had a diagnosis or symptoms of depression. However, most of these studies did not investigate depression outcomes, and so, whether or not the antidepressants were prescribed to treat depression may not have been perceived as relevant.

The short follow-up times noted in studies were primarily due to study design features. Cross-sectional designs were often used to measure the associations between current antidepressant use and factors of interest. Longitudinal studies that we identified only had short follow-up times intended to capture short-term outcomes.

Finally, the majority of studies were excluded partly due to lack of information regarding the duration of antidepressant treatment. Most of these measured either “any antidepressant prescription” during the follow-up or “current use” at a baseline date. Again, the purpose of these studies was not to assess long-term outcomes of a full course of antidepressant treatment.

### 4.2 Strengths and limitations

This is the first systematic review to our knowledge that focuses on long-term outcomes of antidepressant treatment prescribed for the minimum recommended duration (6+ months) or longer, from observational studies, in individuals with comorbid depression and T2DM. Previous reviews have focused on RCTs, and the maximum follow-up time in these RCTs was limited to 6 months at most. We hoped to identify further relevant research evidence by searching for suitable observational studies, however, we were unable to identify any studies which met our inclusion criteria.

The search terms used by our review were broad, searching seven databases to provide a wide range of coverage; this resulted in a large number (14,389) of abstracts being screened. In addition, as the review was also exploratory, the range of potential outcomes we aimed to include were wide-ranging.

Although our search terms were broad, our inclusion/exclusion criteria for depression, T2DM, 6 month minimum duration of antidepressant treatment, and overall follow-up time, were strict in order to identify high quality robust studies, resulting in the exclusion of 63 studies at full text stage. We required precise case definitions for both T2DM and depression, however, if we had not done this, studies would have been included that were not specific to our patient group of individuals with comorbid depression and T2DM. While terms for diabetes type in clinical practice have historically been varied, the relationship between T2DM and depression is distinct [1], and so, specific focus on this patient group is required. Ultimately, there is a lack of relevant research in this large and growing patient group.

We also required a minimum of 6 months antidepressant treatment duration, based on treatment guidelines for antidepressants to be effective [6]. We deemed this criteria to be essential, as our outcomes of interest were long-term, rather than focussing on initial response to treatment. Furthermore, “any” or “current” antidepressant use, based on a single antidepressant prescription, would include individuals who have been prescribed an antidepressant once, without ever having taken it. We aimed to assess people who had received a full treatment course for depression.

### 4.3 Conclusions and Implications

Despite potentially high numbers of individuals with comorbid depression and T2DM being treated with antidepressants, the long-term impact of antidepressant treatment in this patient group is unknown. Focused research is urgently required on the long-term impact of antidepressant treatment, on both physical and mental health outcomes, in individuals with comorbid depression and T2DM.

## Data Availability

This is a systematic review - no studies met inclusion criteria and so the only data included are reasons for study exclusion, contained in the manuscript.

